# Quality of primary healthcare for chronic diseases in low-resource settings: evidence from a mixed methods study in rural China

**DOI:** 10.1101/2024.05.10.24307203

**Authors:** Mingyue Li, Xiaotian Zhang, Haoqing Tang, Huixian Zheng, Ren Long, Xiaoran Cheng, Haozhe Cheng, Jiajia Dong, Xiaohui Wang, Xiaoyan Zhang, Pascal Geldsetzer, Xiaoyun Liu

## Abstract

**Background:** There is a paucity of evidence regarding the definition of the quality of primary healthcare (PHC) in China. This study aims to develop a modified conceptual framework PHC quality based on the context of rural China and evaluate the PHC quality for chronic diseases in rural areas.

**Methods:** This mixed-methods study, involving a patient survey, a provider survey and chart abstraction, and second-hand registered data, was set in three low-resource counties in rural China from 2021 to 2022. Rural patients with hypertension or type 2 diabetes, health care workers providing care on hypertension or diabetes were involved. Standardized PHC quality score was generated by arithmetic means or Rasch models of Item Response Theory.

**Results:** A modified PHC quality framework was presented. High-quality PHC for chronic diseases encompasses three core domains: a competent PHC system (comprehensiveness, accessibility, continuity, and coordination), effective clinical care (assessment, diagnosis, treatment, disease management, and provider competence), and positive user experience (information sharing, shared decision-making, respect for patients’ preferences, and family-centeredness). This study included 1355 patients, 333 healthcare providers and 2203 medical records. Ranging from 0 (the worst) to 1 (the best), the average quality score for PHC system was 0.718, with 0.887 for comprehensiveness, 0.781 for accessibility, 0.489 for continuity, 0.714 for coordination. For clinical care, average quality was 0.773 for disease assessment, 0.768 for diagnosis, 0.677 for treatment, 0.777 for disease management, and 0.314 for provider competence. The average quality for user experience was 0.727, with 0.933 for information sharing, 0.657 for shared decision-making, 0.936 for respect for patients’ preferences, and 0.382 for family-centeredness. The differences in quality among population subgroups, although statistically significant, were small.

**Conclusion:** The PHC quality in rural China has showed strengths and limitations. We identified large gaps in continuity of care, treatment, provider competence, family-centeredness, and shared decision-making. Policymakers should invest more effort in addressing these gaps to improve PHC quality.

## Introduction

Primary healthcare (PHC) is the cornerstone of any health systems. The 2018 Declaration of Astana reaffirmed the fundamental role of PHC in enhancing people’s physical and mental health as well as social well-being [1]. It is universally acknowledged that PHC creates the foundation to achieve universal health coverage (UHC) and the health-related Sustainable Development Goals (SDGs). The underlying assumption is that PHC has adequate quality to optimize health. However, this assumption is not always valid, especially in low-income and middle-income regions. Studies have shown that widespread gaps still exist in PHC quality [2] [3], such as undertreatment and poor management of non-communicable diseases [2], overuse of antibiotics [4] and low coordination of services [5]. Poor PHC quality can jeopardize trust, leading people to bypass PHC and seek expensive specialty care.

PHC quality is strongly correlated to the services provided. Due to population aging and shifts in lifestyles, the burden of chronic diseases has rapidly increased, notably hypertension-related diseases and diabetes mellitus. According to a 2017 study in 154 countries, the annual deaths associated with systolic blood pressure of 140 mmHg or higher has increased from 97.9 to 106.3 per 100,000 population from 1990 to 2015[6]. There are 529 million people living with diabetes worldwide in 2021, and the disability-adjusted life-years (DALY) count has increased by 189.8% since 1990[7]. Growing chronic diseases burden have resulted in changes in PHC delivery and health services. In China, multiple national strategies have been in place to strengthen PHC, such as building medical alliances, launching family doctor contracting services program and integrating medical and preventive services[8]. Changes in delivery and services will have profound impact on how we define and measure quality. For example, the transition of care from hospitals to community settings for chronic diseases (changes in delivery) and shift towards integrating prevention services, such as early screening for hypertension, into routine clinical care (changes in services) will impact PHC quality.

PHC quality is closely associated with contextual factors, including disparities in health systems, economic power, and political backgrounds, alongside the spectrum and burden of diseases. Applying a uniform PHC quality framework without accounting for these variations may lead to incongruities. Typically, researchers modify the general framework to suit local context before implementation. For example, Rezapour et al. developed an Iranian PHC quality framework based on WHO framework and other frameworks, ensuring alignment with Iran’s health system[9].

In theory, PHC is well positioned to respond to chronic diseases if it promotes first-contact and a broad range of basic care to all people[10]. However, PHC in rural areas is often understaffed, placing healthcare workers under strain to deal with the growing disease burden[11]. Constrained resources and insufficient external support also undermine the job performance and productivity of healthcare workers[12]. PHC workers in rural areas often have low medical qualification, which subsequently leads to low-quality services[13] [14]. Studies using incognito standardized patients (also referred to as audit study) found that rural PHC clinicians can only complete 18% of a checklist comprising recommended questions and exams in China[15]. Studies evaluating PHC quality in rural areas have mainly relied on standardized patients, vignettes, or subjective measurement of patient experiences [16] [17]. These methods primarily reflect the quality of episodic care. The unique advantages of PHC in responding to chronic diseases, by comprehensively and continuously meeting most basic healthcare needs at low costs, are overlooked in these methods.

Most research about PHC quality has been conducted in high-income countries, while the respective metrics are frequently unavailable in rural areas. Global initiatives have been actively advancing studies on PHC quality in low-income and middle-income countries [18] [3] [19]. We also analyzed and compared current PHC quality frameworks in a systematic review[20]. These frameworks have established a solid knowledge base for understanding PHC quality.

Notably, the mapping of PHC indicators to High-Quality Health System (HQSS) framework [3] and Primary Health Care Performance Initiative (PHCPI) framework are two major efforts[21]. As a primary source of information, we draw on the HQSS framework. It has garnered widespread attention in defining and assessing PHC quality in low-income and middle-income countries. The HQSS includes three domains: 1) competent systems (safety, prevention and detection, continuity and integration, population-health management, and timely action), 2) evidence-based care (technical quality indices for key PHC services), 3) positive user experience (patient focus, clear communication) [3, 22]. Furthermore, this framework shed lights on the “black box” (the process of care) of PHC quality because they offer a thorough and practical depiction of PHC quality. As a second source, we use the PHCPI framework, a collaborative effort involving the Bill & Melinda Gates Foundation, the World Bank, WHO, and other partners[18, 21]. PHCPI offers a comprehensive evaluation of the overall performance of PHC system, with a specific emphasis on service delivery. Within the service delivery section of PHCPI, key dimensions include population health management, facility organization and management, access, availability of effective PHC services, and high-quality PHC. PHCPI emphasizes essential structural contents tailored for low-income and middle-income countries, encompassing information systems and facility management. Our research aligns with the quality focus delineated in the service delivery section of PHCPI, with a particular emphasis on the distinctive functions of PHC.

This study contributes to the existing literature along the following two dimensions. First, we aim to improve the applicability and relevance of the conceptual framework on PHC quality by incorporating insights from recent global frameworks of PHC quality. Second, we aim to assess PHC quality for chronic diseases in rural China based on a modified framework, with the objective of identifying areas requiring improvement. Hypertension and diabetes were used as tracer conditions. In sum, our findings add knowledge on understanding and enhancing PHC quality for chronic diseases, not only in rural China but also in other low-income and middle-income countries that experience transformative phases in their health systems.

## Methods

### The settings

This cross-sectional study was conducted in three counties of three provinces in western and central China, Hubei, Henan, and Shanxi Province. The socioeconomic status in the three counties was all below the national average – the annual per capita disposable income of rural residents was $2006, $2166, and $1940 in the project counties Hubei, Henan, and Shanxi Province (exchange rate: 1 CNY=0.14 USD) in 2022, compared to $2840 in China. The health resources were also below the national average level – the number of doctors per thousand population was 2.88, 2.38, and 2.72 for the three project counties in 2022, compared to 2.90 in China (table A3).

In rural China, primary care is predominantly delivered by village clinics, township health centers and county hospitals. Most village clinics are privately owned, with some being affiliated with township health centers. Township health centers are public institutions and receive full government subsidies, while county hospitals, also public institutions, are partially subsidized by government funding. County hospitals traditionally provided secondary care, but as more people seeking hospital outpatient care as first-contact care, county hospitals are now playing the role of PHC. Hospitals providing PHC is not uncommon. A survey in 14 countries showed that conditional on people who have usual source of care, 35.6% reported that their usual source of care is in hospitals [23].

China’s rural PHC system is undergoing reforms, with the most notable being medical alliance system that started in 2015 [24]. County hospitals and PHC facilities build networks of care, with or without unified administrative management. County hospitals and PHC facilities are assigned with clear roles and responsibilities. The reform aims to build a patient-centered dual referral pathway alongside the care continuum[8]. In the three project counties, county A in Shanxi province has built one medical alliance including all PHC facilities with a unified administrative system (administration, personnel, business, funds, medical devices, and performance evaluation etc.). County B in Henan province and County C in Hubei province have both built two medical alliances within each county, in collaboration with several PHC facilities, each led by a county hospital.

PHC facilities have transitioned from solely focusing on clinical care to expanding to public health services. To respond to challenges of chronic diseases, China launched the National Basic Public Health Service Program in 2009. PHC facilities started to deliver a comprehensive package of public health services to all residents free-of-charge, encompassing vaccination, health examinations, screening, health management and health education. A key component of the service package is the registration and management of patients with hypertension or type 2 diabetes. The funding for the program is invested from both central and local governments and is allocated to PHC facilities based on capitation [25].

The concept of participants involvement translated to the study design and execution phases of the research. The development of the research question was based on public concerns. Patients and doctors were recruited to the study, and the study design was explained in detail. We worked with investigators from communities, and they reviewed the questionnaires first and gave feedback. We disseminated the study results by public policy briefs.

### Conceptual framework of PHC quality

Drawing on HQSS and PHCPI, we updated the conceptual framework for evaluating the PHC quality for chronic diseases in rural China. High-quality PHC for chronic diseases should underpin three core domains: a competent PHC system, effective clinical care, and positive user experience (Fig 1). We added the provider competence from the PHCPI to the clinical care sub-domain because providers in rural China have been long criticized to have insufficient ability and medical knowledge [2]. We included disease management as a sub-domain due to our focus on chronic diseases and the increased time spent on disease management by PHC providers after the National Basic Public Health Service Program implementation. We simplified sub-domains for user experiences compared with the HQSS, because some of them are not suitable for rural China’s context. For example, discrimination or social stigma may be associated with certain chronic diseases (such as hepatitis B, mental illness, tuberculosis, or HIV infection) in rural China [26], but it is not a primary concern for hypertension or diabetes. While we recognized that inputs and outcomes were also highly important, this study identified quality more as a concept of process. The building blocks of health system inputs, including drugs and supplies, facility infrastructure, health workforce, health financing, and information systems, were fundamental in building a competent PHC. PHC without positive outcomes, including health outcomes, financial burden, equity, responsiveness, and resilience, is ineffective.

**Fig 1.**
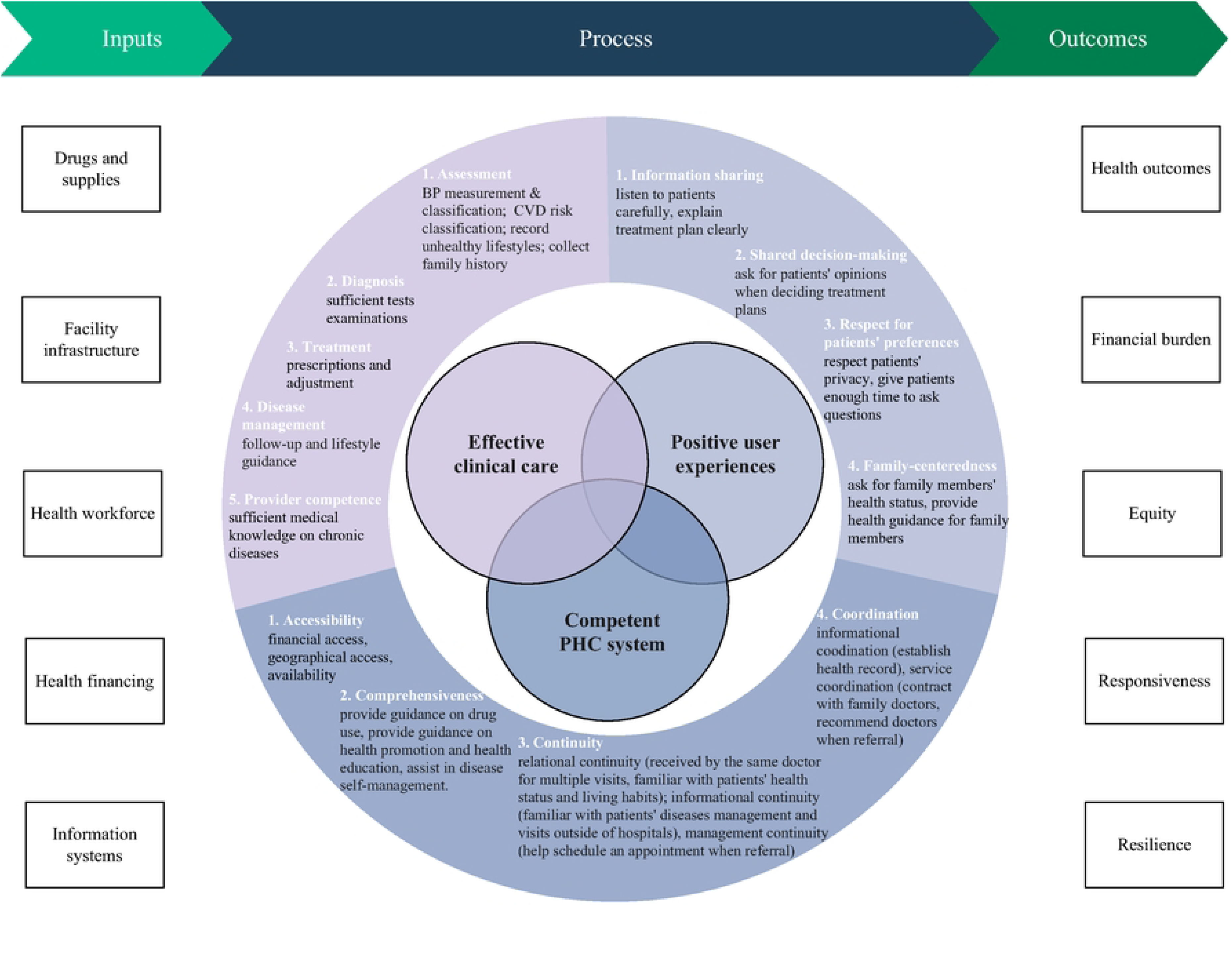
The modified conceptual framework for the primary healthcare quality for chronic diseases in rural China

The definitions for quality domains and sub-domains are presented in Table 1. Quality for PHC system focused on the unique functions of PHC compared to specialty care. Accessibility, comprehensiveness, continuity, and coordination were universally recognized as essential to achieve an effective PHC system. Quality for clinical care focused on the continuum of patient journey for chronic diseases which refers to the degree to which clinical processes conform to existing evidence and/or widely accepted standards[3, 27]. For chronic diseases, the continuum of care covers disease assessment, diagnosis, treatment and disease management, and provider competence that ensured an effective care continuum. Quality for user experience refers to information sharing, shared decision-making, respect for patients preferences and family-centeredness [28].

**Table 1.**
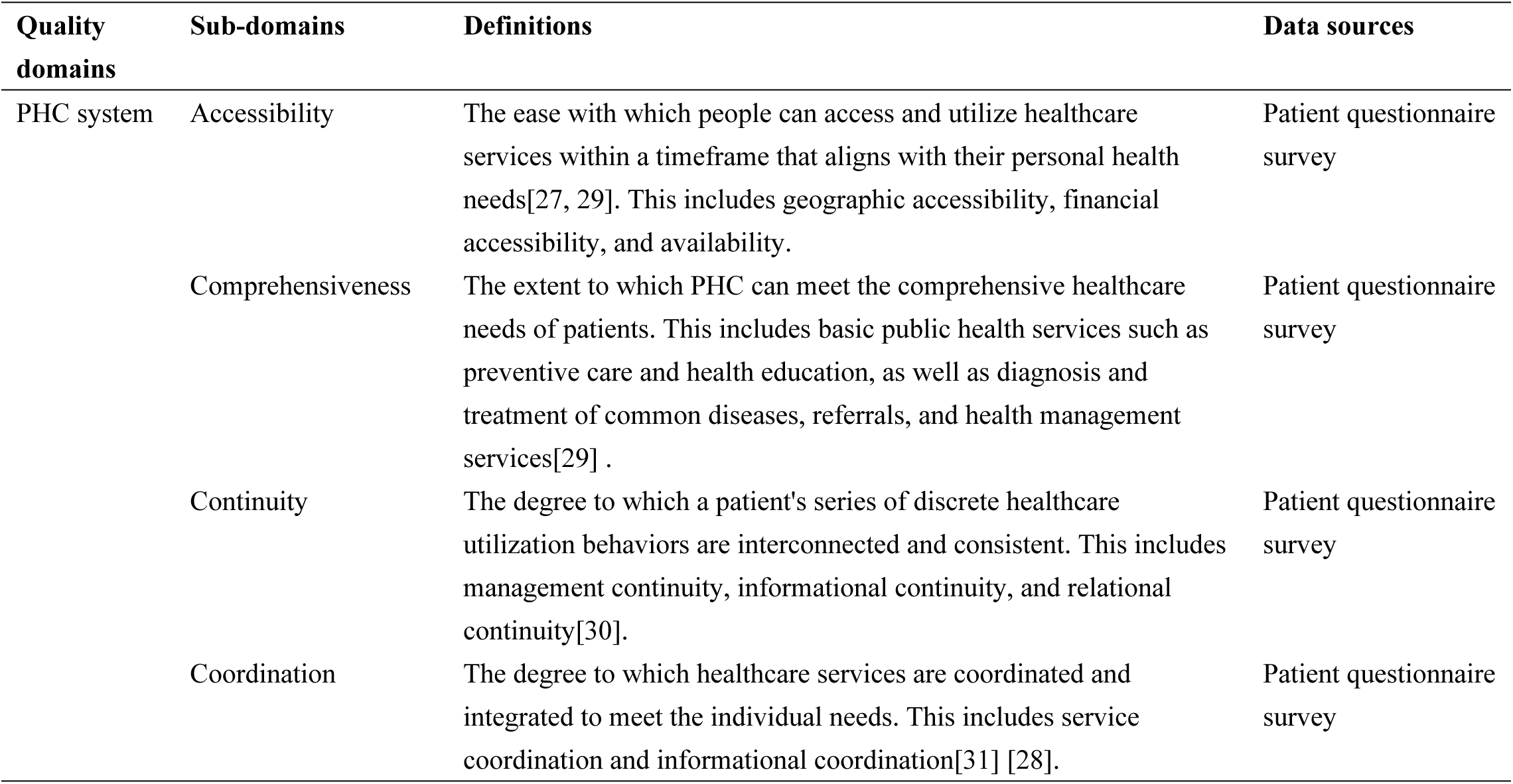
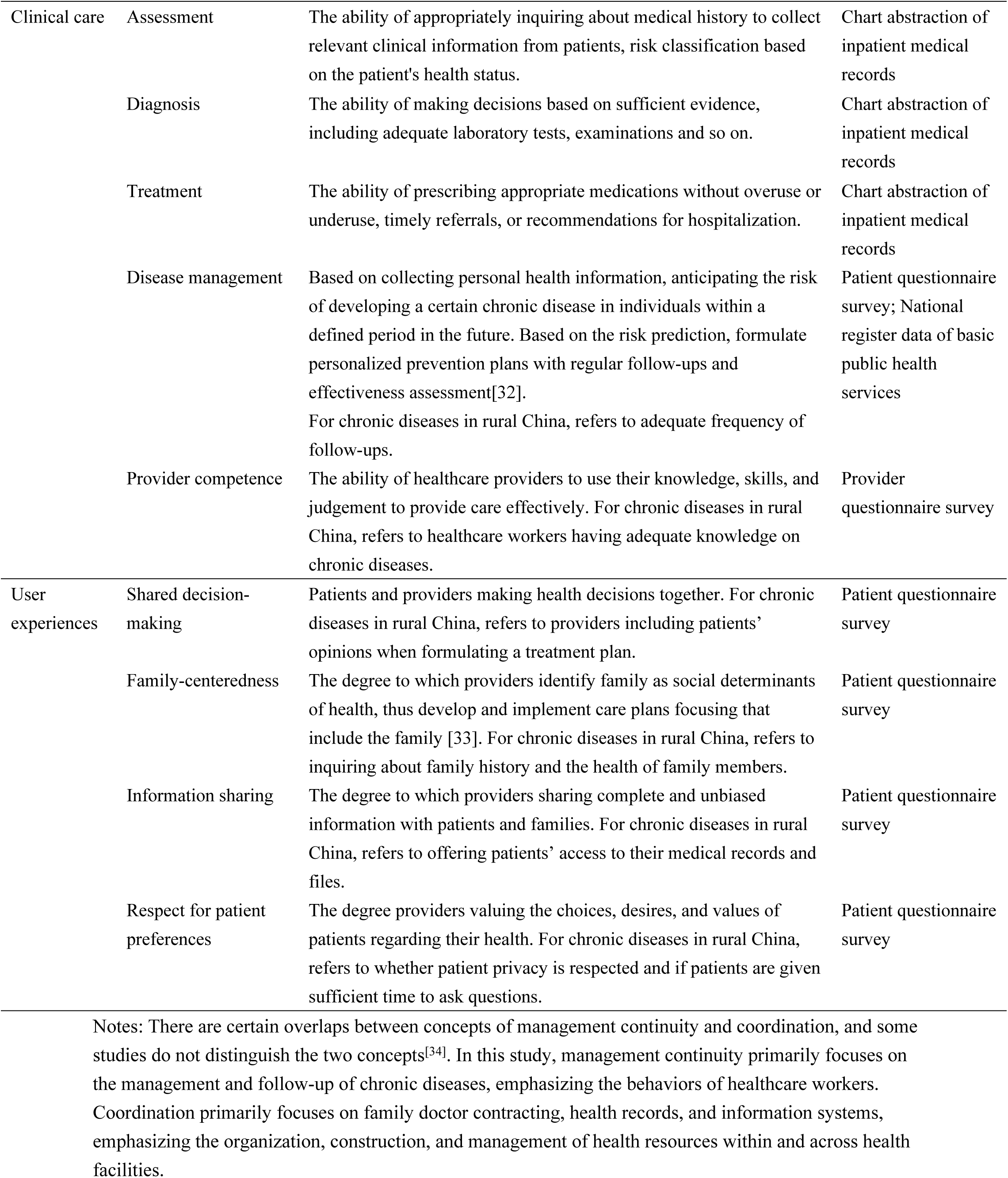
Definitions and data sources for quality domains and subdomains.

### Data source and participants

Multiple data sources including patient questionnaire survey, provider questionnaire survey, medical records of health facilities, and register data from governments were used to evaluate PHC quality (Table 1). Hubei, Henan, and Shanxi provinces that locate in less-developed western and central areas were selected based on willingness of cooperation and logistic considerations. Three low-resource counties and nine towns were selected. The administrative structure in China is strictly hierarchical, consisting of province/municipality at the first level, and a county, a township, or a village in a rural area.

We conducted a patient questionnaire survey to collect data on PHC system and user experiences. We used a two-stage stratified cluster random sampling. The strata were regions (county A in Hubei, county B in Henan, and county C in Shanxi) and villages. In the first stage, a random sample of 9 towns were selected whereby 3 towns were selected in each county. In the second stage, 900 hypertensive patients and 450 diabetic patients were selected whereby 300 hypertensive patients and 150 diabetic patients were selected in each county. Registration dates before 07/30/2022 were included. Because patients were not randomly registered but based on the dates of disease detection, and newly detected patients typically exhibit milder symptoms, we used a systematic random sampling approach of patients’ villages instead of selecting individual patients. This method enabled us to include patients with diverse levels of severity. All individuals with hypertension or diabetes living in the selected villages were recruited between 07/10/2022 and 07/27/2022. The questionnaire can be found in the appendix. We also conducted a provider survey to collect information on provider competence.

Scenarios of patients with varied severity of hypertension or diabetes were presented to providers. We used a cluster sampling approach, and all the health care workers providing care on hypertension or diabetes (village doctors, general practitioners (GP), public health doctors, internists) in the three county hospitals (n=50), nine towns (n=76) and all villages (n=199) were recruited between 07/11/2022 and 07/27/2022. The response rate for providers was 94.0% in county hospitals, 84.2% in township health centers and 89.9% in village clinics. The questionnaire can be found in the appendix.

We reviewed and abstracted medical records using pre-determined checklists to collect data on disease assessment, diagnosis and treatment, a method known as chart abstraction, which is widely used to evaluate quality of care[17, 35]. Basically, we collected detailed information on clinical processes, such as whether performing glycated hemoglobin test for diabetic patients or whether classifying patients based on blood pressure level. The detailed checklists can be found in the appendix. Patients with primary discharge diagnosis of hypertension, diabetes, or related complications from 01/01/2020 to 06/30/2022 were included. Patients with other primary diagnosis (i.e. poisoning) but with a history of hypertension, diabetes or related complications were excluded. Data were primarily abstracted from inpatient medical records, including both paper and electronic records. Other electronic health systems were also used for supplementary information, including the Hospital Information System (HIS) system, the imaging system, and the laboratory system. Clinical experts with pre-trained investigators reviewed medical records, abstracted information, and filled up the checklists. All checklists were double-checked at the end of day by investigators and clinical experts. The medical records were accessed between 07/11/2023 and 07/27/2023 for research purposes, and no identifiable individual information was collected.

Health records and follow-up visits for chronic diseases are registered in the national basic public health services system. The anonymized data on hypertension/diabetes follow-up visits were exported from the registry. The data contained time and blood pressure/glycated hemoglobin measurement for each follow-up visit. Detailed description of indicators for each domain and sub-domain are presented in the appendix.

### Calculation of quality scores

Five quality scores were calculated for three domains of PHC quality at the patient or provider level. Due to various data sources, clinical care is reflected by separate sub-domain scores. The score of provider competence was calculated at the provider level, while all other scores were calculated at the patient level. The list of indicators for each domain and sub- domain is reported in the appendix.

All PHC quality domains were composed of binary (ie, 0 or 1) indicators and were then summarized into domain score or sub-domain score ranging from 0 (lowest) to 1 (highest). Scores above 0.7 were considered as favorable or optimal. Different strategies were used for summarizing quality scores due to data structure and sources: 1) the arithmetic mean was calculated to get the quality scores for PHC system, disease management, and user experience; and 2) Item Response Theory (IRT) models were used to get quality scores for clinical care and provider competence. IRT has been widely used in educational and psychological testing for studying the relationship between participants’ latent characteristics (such as competence, cognition) and their response to a set of questions. IRT has also been used in studies on PHC quality. For example, Das et al. also used IRT to analyze PHC quality in India in 2004[36]. We used the Rasch model (also referred to as one parameter IRT model) to calculate quality scores for clinical care and provider competence. For the Rasch model (1), P is the probability of person j’s correct response to question i, and θ is the provider competence. The hypothesis was that questions vary only in difficulty (b_i_). The Rasch model can be transformed into (2), i.e. a typical logistic model, that can be estimated by maximum likelihood method:

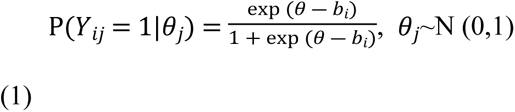

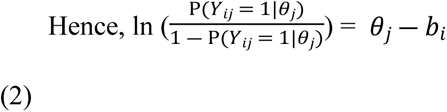

### Statistical analysis

We reported the levels of quality scores for each quality domain and sub-domains, and then reported the distribution of quality scores by socioeconomic characteristics. We assessed gender, age, marriage, education, household income group, and medical insurance. We also conducted a series of sensitivity analyses to test how robust our findings were to modeling assumptions. Cronbach’s alpha was used to examine the reliability of the summative score (appendix). Since no summative score was built for clinical care domain, we used multivariable linear regression models to further examine the association between provider characteristics and provider competence. As predictors, we assessed working places, gender, education, having permanent posts, having passed medical licensing exam, job title, administrative roles, and job satisfaction. No sampling weights was used, and standard errors were clustered at the level of regions (towns). Missing data (unanswered questions) were imputed using multiple chained equations. This study follows the STROBE checklist for observational cross-sectional studies. P<0.05 was considered as statistically significant. All statistical analyses were conducted in Stata V.17.0 (Stata Corp LP).

## Results

### Characteristics of the study sample

In total, this study included 1355 patients, 333 providers and 2203 medical records (Table 2). Patients in the patient survey were on average 65.8 years’ old, and 20.4% of the patients had a comorbidity of hypertension and diabetes. Providers came primarily from village clinics (53.7%). 77.9% of the inpatient medical records were from county hospitals. The average age for inpatients was 64.3 years’ old.

**Table 2.** Characteristics of the study sample.

|  | **Patients** | **Providers** | **Medical records** |
| --- | --- | --- | --- |
| Sample size (N) | 1355 | 333 | 2203 |
| Regions, N (column %) |  |  |  |
| Hubei | 420 (31.0) | 132 (39.6) | 794 (36.0) |
| Henan | 466 (34.4) | 143 (42.9) | 726 (33.0) |
| Shanxi | 469 (34.6) | 58 (17.4) | 683 (31.0) |
| Age, Mean (SD) | 65.8 (9.4) | 41.3 (10.5) | 64.3 (12.2) |
| Gender, N (column %) |  |  |  |
| Female | 845 (62.4) | 162 (48.6%) | 1085 (49.3%) |
| Male | 510 (37.6) | 171 (51.4%) | 1118 (50.7%) |
| Health facilities, N (column %) |  |  |  |
| Village clinics | NA | 179 (53.7) | NA |
| Township Health Centers | NA | 107 (32.1) | 487 (22.1) |
| County hospitals | NA | 47 (14.1) | 1716 (77.9) |
| Survey year | 2022 | 2022 | 2021-2022 |
| Diseases, N (column %) |  |  |  |
| Hypertension | 901 (66.5) | NA | 850 (38.6) |
| Diabetes | 178 (13.1) | NA | 792 (36.0) |
| Comorbidity | 276 (20.4) | NA | 561 (25.5) |

*NA, not applicable. Village clinics in China do not provide inpatient services.*

Till now, there are two nationally representative surveys on hypertension and diabetes in China[37, 38]. In the national hypertension survey, 57.6% of the hypertensive patients were female in 2017; in the national diabetes survey, 50.0% of the diabetic patients were women, and the median age was 51.3. Our study only focused on patients from low-resource rural areas, which may not be comparable to the nationally representative surveys. Patients in our study comprised more women (62.4%) and were older than national representative samples.

### Levels of PHC quality

Fig 2 shows the PHC quality score for different domains and sub-domains. The overall PHC system score was 0.718 on average, and user experience total score was 0.727 on average. Within the PHC system quality domain, comprehensiveness received the highest score (0.887), while continuity scored the lowest (0.489).

**Fig 2.**
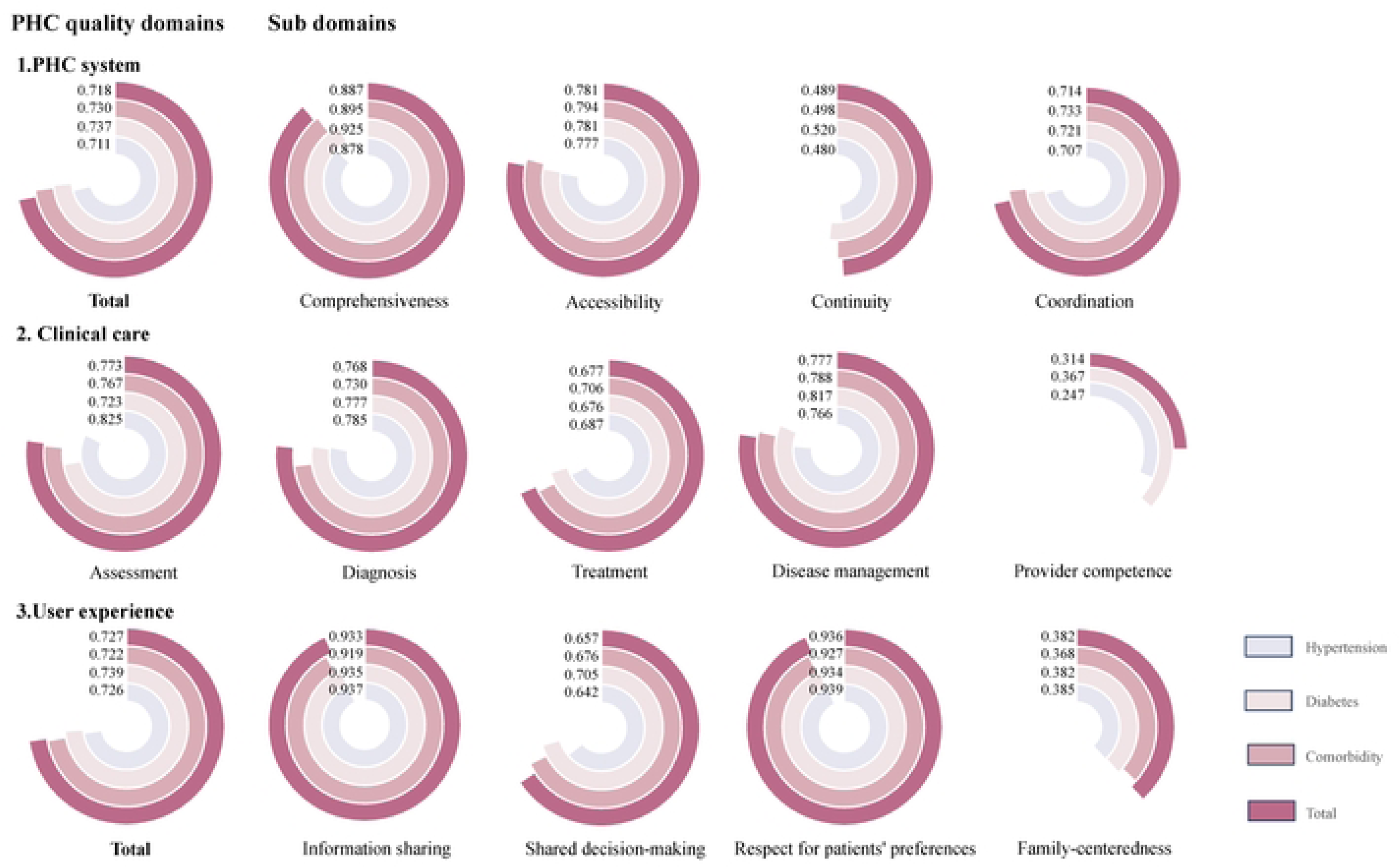
Average quality domain and sub-domain score of PHC for hypertension, diabetes, and comorbidity. For quality score, 0 represents the lowest quality, and 1 represents the highest quality.

Clinical care score was displayed by sub-domains due to disparate data sources. Assessment (0.773), diagnosis (0.768), and disease management (0.777) demonstrated similar and favorable scores (scoring above 0.7 out of 1), whereas provider competence scored lower (0.314).

Within the user experience quality domain, information sharing (0.933) and respect for patients’ preferences (0.936) scored higher compared to shared decision-making (0.657) and family-centeredness (0.382).

The quality scores for diabetes were generally higher (0.737 for diabetes vs 0.711 for hypertension in PHC system, 0.739 for diabetes vs 0.726 for hypertension in user experience), but the variations in the quality scores across diabetes, hypertension, and comorbidity were small.

## Distribution of PHC quality

Fig 3 presents the PHC quality score and 95% confidence intervals by socioeconomic status, and Fig A1 presents the results on statistical tests (Appendix, Fig A1). The scores for comprehensiveness of care were higher for women (0.90 vs 0.87, P=0.033), while the opposite was observed for scores in coordination (0.70 vs 0.74, P=0.007). The younger patients (<50) showed higher quality scores in most quality dimensions compared to the elder groups (≥70), but similar scores in continuity (0.50 vs 0.50). Married patients revealed higher scores in accessibility (0.79 vs 0.76, P<0.001) and family-centeredness (0.40 vs 0.30, P<0.001).

**Fig 3.**
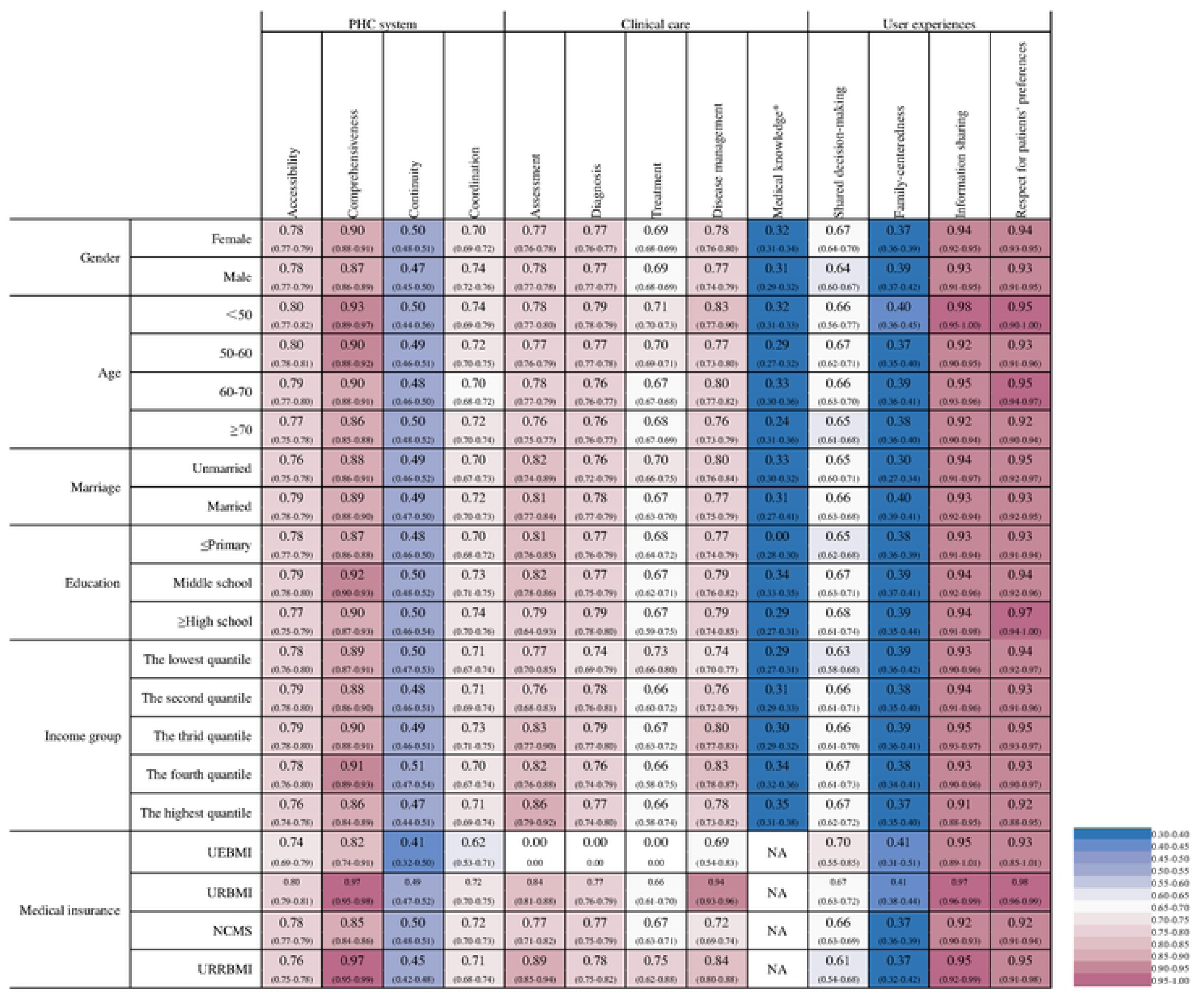
The distribution of average PHC quality score by social-economic characteristics (95% CI). The distribution of quality score for assessment, diagnoses, and treatment in marriage, education, income group, and medical insurance were based on matched data between chart abstraction and patient survey; Quality score for medical knowledge is at provider level rather than patient level. NA, not applicable, meaning no observations in that category. UEBMI, Urban Employee Basic Medical Insurance. URBMI, Urban Resident Basic Medical Insurance. NCMS, New Rural Cooperative Medical Scheme. URRBMI, Urban and Rural Resident Basic Medical Insurance

The scores for comprehensiveness of care were higher in patients with higher education level (high school) compared to those with primary school (0.90 vs 0.87, P<0.001), but lower compared to middle school (0.90 vs 0.92, P<0.001). Lower income groups showed lower quality scores in diagnosis (0.74 for the lowest income group vs 0.77 for the highest income group, P=0.045), disease management (0.74 vs 0.78, P=0.002), but higher scores in accessibility (0.78 vs 0.76, P<0.001), comprehensiveness (0.89 vs 0.86, P=0.003) and other dimensions in quality of user experience.

We conducted reliability checks for quality domains of PHC system, user experience and provider competence, and the respective Cronbach’s α was 0.583 and 0.711 (appendix). As for clinical care, we followed Chinese clinical guidelines for hypertension and diabetes, National Basic Public Health Service Standards, and consulted expert opinions. We conducted sensitivity analyses to examine the associations between a number of factors and provider competence. Compared with providers at village clinics, providers at township health centers and county hospitals had significantly better competence. Providers at county hospitals had 26% higher competence than those at village clinics (P=0.005). This association was consistent across models (Table 3). Providers with permanent posts had 10% better competence than those without (P=0.039). Education was not statistically significant because it has been largely explained by working places - 75% of providers at village clinics had a high school or below education while all providers at county hospital and 92% providers at township health centers had a college or above degree.

**Table 3.**
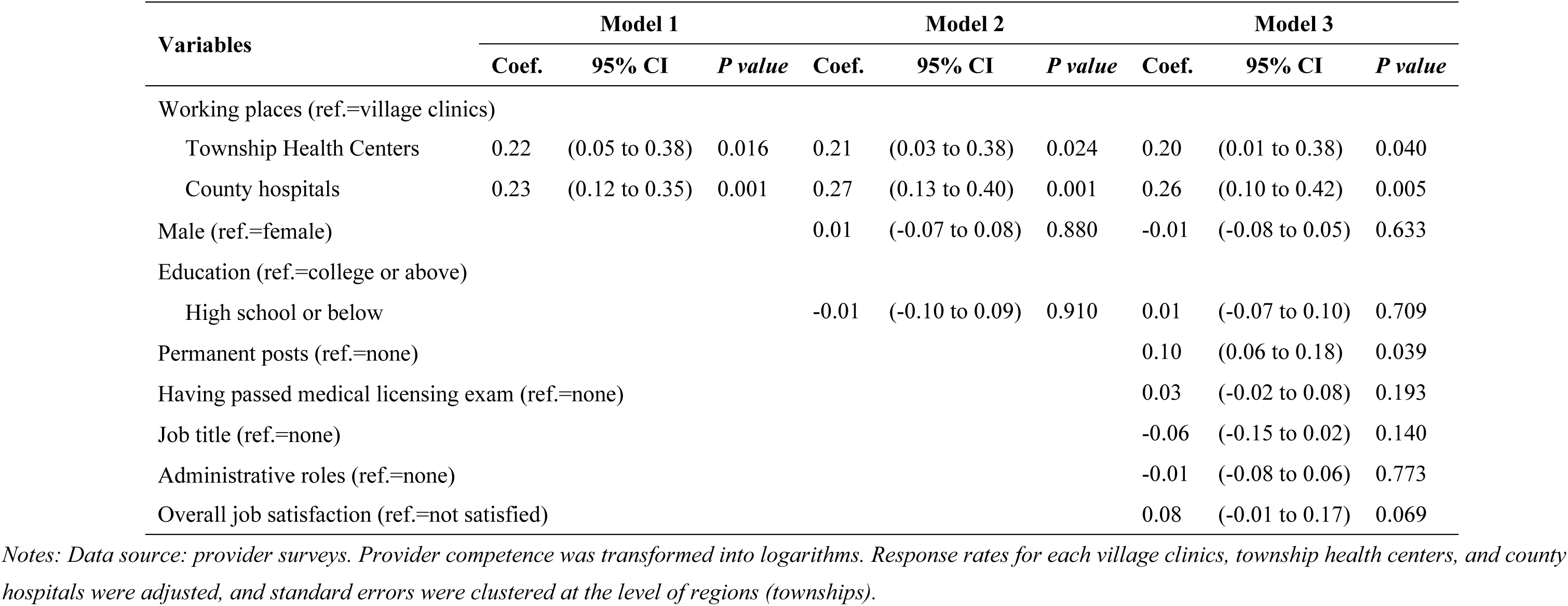
The associations between provider’s characteristics and provider competence.

## Discussion

This study modified the PHC quality framework to the context of rural China and evaluated the PHC quality for chronic diseases drawing on multiple data sources. We would like to highlight the following findings. With a focus on the "process" dimension, PHC quality for chronic diseases can be divided into three domains: quality of the PHC system, quality of clinical care, and quality of user experience. PHC quality in rural China reveals a nuanced situation marked by both strengths and shortcomings. Most quality sub-domains demonstrate favorable levels (scoring more than 0.7 out of 1); while continuity of care, treatment, provider competence, family-centeredness and shared decision-making have significant deficiencies. The differences among population subgroups in PHC quality, although statistically significant, are small in relation to the gap between actual and optimal PHC quality. These results reflect the systematic nature of low-quality problems in low-resource areas. The primary challenge is to enhance the generally suboptimal PHC quality at the aggregate level.

The PHC system exhibits a satisfactory quality score with respect to accessibility, comprehensiveness, and coordination, but a remarkably low score for continuity. We consider several possible explanations for this result. The achievements in accessibility reflected China’s efforts in the past two decades. Providing its citizens with equitable and affordable access to basic health care and financial protection has been set as priority in national policies[39]. China has built a national social health insurance scheme covering nearly all population, and catastrophic medical insurance and medical aid to strengthen financial protection for low-resource populations since 2009[40]. Lacking qualified health workforce in rural areas is a major challenge for improving accessibility , especially in low-resource western and central areas of China. To address this problem, China launched a national compulsory services program to train and to provide qualified general practitioners with a five-year medical training program in these areas since 2010. The program has been implemented for more than 13 years and provides more than 5000 general practitioners each year [12]. China has also invested great efforts in improving comprehensiveness, notably the launch of a comprehensive national public health services program [25]. This program included the establishment of health records for all, health education, elderly care, care for major chronic diseases (including hypertension and type-2 diabetes) and infectious diseases, vaccinations, hygiene monitoring and more[25]. For patients with hypertension or type-2 diabetes, the service package includes screening, monitoring, routine follow-up and customized interventions [41]. PHC facilities provide these basic public health services to residents free of charge. Stable funding based on capitation from central government and local governments ensures the financing of the program. The scope of services provided by PHC has greatly expanded due to this program. A national evaluation study of this program showed that residents were widely satisfied with the service of PHC providers[41]. The achievements in coordination can be attributed to China’s massive investments in building a PHC information system. Health information systems can improve quality of care by improving coordination as a direct mechanism[42]. China’s PHC facilities used to solely rely on paper-based medical records, and now nearly half of the rural facilities are using electronic information systems[41]. There are problems applying information technology in rural areas, for example, low willingness to adopt new information technology[43] and poor record-keeping behaviors[44].

The deficiency in continuity suggests a high fragmentation exists in the chronic care continuum. China’s current health system is criticized for being hospital-centric – the share of outpatient services in PHC facilities decreased from 62% in 2010 to 50% in 2021. Medical resources concentrate in hospitals, leaving PHC facilities weak. Besides, patients do not trust PHC facilities and turn to crowded hospitals for first-contact care[39]. Within the currently fragmented hospital-centric health system, it is difficult to build care continuity for chronic diseases management and control. China has committed to enhance PHC. Reforms including family physician care models and building an integrated delivery system were launched in recent years, but their impact on PHC quality was underexamined[24].

We found that large gaps remained in treatment quality and provider competence, suggesting PHC providers are not prepared for the growing number of chronic patients and complex healthcare needs. Our findings are in line with pre-existing evidence[37, 39, 45]. UHC progress evaluation showed large gaps in non-communicable disease control and management in China as well as in many other low- and middle-income countries[39]. The treatment of chronic diseases is poor in rural China: a national survey estimated that in 2018 only 28.8% rural patients with diabetes were treated (vs 36.2% in urban); only 44.1% patients being treated were controlled (vs 54.1% in urban)[38]. For patients with hypertension, another national survey showed that in 2017 only 28.2% (vs 33.4% in urban) were taking prescribed antihypertensive medications[37]. The PHC quality in urban and rural areas are both unsatisfactory, but the situation is worse in the latter. China’s rural PHC providers have low levels of education and qualifications: in 2021, only 31.3% of the doctors in township health centers have college degree and 17.5% in village clinics [45]. Without sufficient knowledge on chronic disease, it is impossible to improve clinical quality. Hospitals, medical universities, or associations should provide more training programs for PHC providers. Pairing assistance between secondary hospitals and PHC institutions will also benefit PHC providers’ improvement [46].

We found large gaps in shared decision-making and family-centeredness. An understanding of the illness and an active participation in monitoring and treatment are essential for effectively managing chronic conditions. China’s PHC is largely based on ‘doctors’ orders – paternalistic advice with limited information sharing. As detailed in the WHO Innovative Care for Chronic Conditions model, a family-centered approach is important for long-term management of chronic diseases because changing lifestyles is often difficult and needs family support [47]. Involving family members helps PHC doctors to better understand patients’ overall situation and to target social, emotional, and environmental determinants. However, rural PHC doctors lack awareness of family-centeredness and effective interventions in a family-centered way. China’s healthcare system should emphasize the role of family in caring for chronic diseases. Shared decision-making and family-centeredness can be taught through training programs.

Our findings that the PHC quality has significant but small variations in different sub- populations are in line with many published studies. PHC quality varies among patient characteristics and provider characteristics, but till now studies did not identify equity of quality as a primary problem. A study comparing quality of care among sociodemographic groups in the US found that women had 57% quality-of-care score, significantly higher than 52.3% of men (P<0.001) – but the difference is small compared to the general low quality score[48]. Another study in India found that poor and rich people living in the same village received similar quality of care[13]. Studies have explored gender differences in quality of care for decades but reached mixed conclusions [49].

We used multiple methods to assess quality: chart abstraction, patient survey, and vignette through provider survey. Results measured by chart abstraction or vignette may underestimate quality, as a result of incomplete documentation, especially for paper-based documentation, and underreporting information. The materials used in our study were primarily from the electronic health information system, which could minimize recording bias or inaccuracy. Chart abstraction has advantages over other methods of assessing quality – lower cost, more convenient access, and a large sample size. Vignette has proved both efficacy and efficiency in measuring quality. A study comparing different methods measuring quality of care estimated that chart abstraction underreported quality 10.6 percent lower than standardized patients, and 5.4 percent lower than vignette[50]. The low quality in clinical care and provider competence can be partially explained by measurement methods, but the shortfalls is too widespread to be fully attributed to methods.

This study is subject to several limitations. The PHC quality scores are primarily based on surveys, which may be subject to bias due to systematic variations in people’s expectations. As a result, even though the quality score is objectively the same, it may be assessed higher by people with lower expectations. We mainly used hypertension and diabetes as tracer diseases. Excluding other major chronic diseases in China (i.e. cancer, respiratory diseases) could lead to some loss of information. Furthermore, the findings of this study, conducted in three under-resourced counties, may not be generalizable to rural areas in China as a whole. In addition, the data obtained from medical records may differently underrepresent actual quality due to potential incomplete documentation by providers.

## Conclusion

We modified the PHC quality framework for chronic diseases and used it for assessing PHC quality in low-resource settings. The quality for chronic diseases in rural China has showed strengths and limitations. Significant deficiencies were identified in PHC system (continuity of care), clinical care (treatment and provider competence) and use experience (family-centeredness and shared decision-making). Future efforts on improving PHC quality should be directed on these areas.

## Data Availability

The datasets generated and/or analyzed during the current study are not publicly available due to limitations of ethical approval involving the personal data and anonymity but are available from the corresponding author on reasonable request.

## Acknowledgements

The authors would like to thank all project members in Hubei, Henan and Shanxi provinces for their support in carrying out all the surveys. Mingyue Li and Prof. Xiaoyun Liu would like to thank Prof. Xiaolin Wei, Prof. Jinzhu Jia, Dr. He Zhu, Dr. Yao Yao for their invaluable advice in the early planning stages of this study. Mingyue Li would like to thank Dr. Fabian Dehos for his genuine advice in revising the manuscript.

## Supporting information

Appendix

## Notes

### Competing Interest Statement

The authors have declared no competing interest.

### Funding Statement

Yes

### Author Declarations

This study was approved by the Institute Review Board (IRB) of Peking University Health Science Center (IRB00001052-22155). Written informed consent was obtained from all participants and health organizations prior to questionnaire.

## References

1. WHO, Declaration of Astana. 2018.

2. Li, X., et al., Quality of primary health care in China: challenges and recommendations. Lancet, 2020. 395(10239): p. 1802–1812.

3. Macarayan, E.K., et al., Assessment of quality of primary care with facility surveys: a descriptive analysis in ten low-income and middle-income countries. Lancet Glob Health, 2018. 6(11): p. e1176–e1185.

4. Nguyen, N.V., et al., Outpatient antibiotic prescribing for acute respiratory infections in Vietnamese primary care settings by the WHO AWaRe (Access, Watch and Reserve) classification: an analysis using routinely collected electronic prescription data. The Lancet Regional Health - Western Pacific, 2023. 30: p. 100611.

5. Yi, H., et al., The competence of village clinicians in the diagnosis and management of childhood epilepsy in Southwestern China and its determinants: A cross-sectional study. Lancet Reg Health West Pac, 2020. 3: p. 100031.

6. Forouzanfar, M.H., et al., *Global Burden of Hypertension and Systolic Blood Pressure of at Least 110 to 115 mm Hg*, *1990-2015*. JAMA, 2017. 317(2): p. 165–182.

7. GBD 2021 Diabetes Collaborators, *Global, regional, and national burden of diabetes from 1990 to 2021, with projections of prevalence to 2050: a systematic analysis for the Global Burden of Disease Study 2021*. The Lancet, 2023. 402(10397): p. 203–234.

8. Xiong, S., et al., Primary health care system responses to non-communicable disease prevention and control: a scoping review of national policies in Mainland China since the 2009 health reform. Lancet Reg Health West Pac, 2023. 31: p. 100390.

9. Rezapour, R., et al., Developing Iranian primary health care quality framework: a national study. BMC public health, 2019. 19(1): p. 911.

10. Kruk, M.E., G. Nigenda, and F.M. Knaul, Redesigning primary care to tackle the global epidemic of noncommunicable disease. Am J Public Health, 2015. 105(3): p. 431–7.

11. Agyekum, E.O., et al., Networks of care to strengthen primary healthcare in resource constrained settings. BMJ, 2023: p. e071833.

12. Li, M., et al., Supporting and retaining competent primary care workforce in low-resource settings: lessons learned from a prospective cohort study. Fam Med Community Health, 2023. 11(4).

13. Das, J. and A. Mohpal, *Socioeconomic Status And* Quality Of Care In Rural India: New Evidence From Provider And Household Surveys. Health Aff (Millwood), 2016. 35(10): p. 1764–1773.

14. Yi, H., et al., The competence of village clinicians in the diagnosis and treatment of heart disease in rural China: A nationally representative assessment. Lancet Reg Health West Pac, 2020. 2: p. 100026.

15. Sylvia, S., et al., Survey using incognito standardized patients shows poor quality care in China’s rural clinics. Health Policy Plan, 2015. 30(3): p. 322–33.

16. Kwan, A., et al., Use of standardised patients for healthcare quality research in low- and middle-income countries. BMJ Glob Health, 2019. 4(5): p. e001669.

17. Das, J. and J. Hammer, Quality of Primary Care in Low-Income Countries: Facts and Economics. Annual Review of Economics, 2014. 6(1): p. 525–553.

18. Bitton, A., et al., Primary healthcare system performance in low-income and middle-income countries: a scoping review of the evidence from 2010 to 2017. BMJ Glob Health, 2019. 4(Suppl 8): p. e001551.

19. Lewis, T.P., et al., Health service quality in 2929 facilities in six low-income and middle-income countries: a positive deviance analysis. Lancet Glob Health, 2023. 11(6): p. e862–e870.

20. Li, M., et al., Exploring the quality of primary health care: a systematic review (in Chinese). Chinese General Practice, 2024: p. 0-.

21. Veillard, J., et al., Better Measurement for Performance Improvement in Low- and Middle-Income Countries: The Primary Health Care Performance Initiative (PHCPI) Experience of Conceptual Framework Development and Indicator Selection. Milbank Q, 2017. 95(4): p. 836–883.

22. Kruk, M.E., et al., High-quality health systems in the Sustainable Development Goals era: time for a revolution. Lancet Glob Health, 2018. 6(11): p. e1196–e1252.

23. Croke, K., et al., Primary health care in practice: usual source of care and health system performance across 14 countries. The Lancet Global Health, 2024. 12(1): p. e134–e144.

24. Cai, C., et al., Health and health system impacts of China’s comprehensive primary healthcare reforms: a systematic review. Health Policy Plan, 2023. 38(9): p. 1064–1078.

25. Yuan, B., et al., Strengthening public health services to achieve universal health coverage in China. BMJ, 2019. 365: p. l2358.

26. Deng, Y., et al., Mental health-related stigma and attitudes toward patient care among providers of mental health services in a rural Chinese county. International Journal of Social Psychiatry, 2021. 68(3): p. 610–618.

27. Lévesque, J.F., et al., Mapping the coverage of attributes in validated instruments that evaluate primary healthcare from the patient perspective. BMC Fam Pract, 2012. 13: p. 20.

28. O’Malley, A.S., et al., Disentangling the linkage of primary care features to patient outcomes: a review of current literature, data sources, and measurement needs. Journal of general internal medicine, 2015. 30(3): p. 576–585.

29. Haggerty, J., et al., Operational definitions of attributes of primary health care: Consensus among Canadian experts. Ann Fam Med, 2007. 5(4): p. 336–344.

30. Starfield, B., L. Shi, and J. Macinko, Contribution of primary care to health systems and health. Milbank Q, 2005. 83(3): p. 457–502.

31. Kringos, D.S., et al., The European primary care monitor: structure, process and outcome indicators. BMC Fam Pract, 2010. 11: p. 81.

32. Chinese Health Association, Good Health Management Practice for Chronic Diseases (T/CHAA 007-2019) (in Chinese). Chin J Epidemiol, 2020. 14(1): p. 12–14.

33. National Academies of Sciences, E. and Medicine, Implementing high-quality primary care: rebuilding the foundation of health care. 2021.

34. Lévesque, J.F., et al., Canadian Experts’ Views on the Importance of Attributes within Professional and Community-Oriented Primary Healthcare Models. Healthc Policy, 2011. 7(Spec Issue): p. 21-30.

35. World Health Organization, et al., Improving healthcare quality in Europe: Characteristics, effectiveness and implementation of different strategies. 2019: World Health Organization. Regional Office for Europe.

36. Das, J. and J. Hammer, Which doctor? Combining vignettes and item response to measure clinical competence. J Dev Econ, 2005. 78(2): p. 348–383.

37. Lu, J., et al., Prevalence, awareness, treatment, and control of hypertension in China: data from 1·7 million adults in a population-based screening study (China PEACE Million Persons Project). Lancet, 2017. 390(10112): p. 2549–2558.

38. Wang, L., et al., *Prevalence and Treatment of Diabetes in China*, *2013-2018*. JAMA, 2021. 326(24): p. 2498–2506.

39. Yip, W., et al., Universal health coverage in China part 1: progress and gaps. The Lancet Public Health, 2023. 8(12): p. e1025–e1034.

40. World Health Organization Regional Office for the Western Pacific, Progress towards universal health coverage: Monitoring financial protection in the Western Pacific Region. 2023: Manila.

41. Xiong, S., et al., Factors associated with the uptake of national essential public health service package for hypertension and type-2 diabetes management in China’s primary health care system: a mixed-methods study. The Lancet Regional Health - Western Pacific, 2023. 31: p. 100664.

42. Atasoy, H., B.N. Greenwood, and J.S. McCullough, The Digitization of Patient Care: A Review of the Effects of Electronic Health Records on Health Care Quality and Utilization. Annual Review of Public Health, 2019. 40(1): p. 487–500.

43. Xia, Z., et al., Perceived Value of Electronic Medical Records in Community Health Services: A National Cross-Sectional Survey of Primary Care Workers in Mainland China. Int J Environ Res Public Health, 2020. 17(22).

44. Kwiatkowska, R., et al., Patients without records and records without patients: review of patient records in primary care and implications for surveillance of antibiotic prescribing in rural China. BMC Health Services Research, 2020. 20(1).

45. National Health Commission, China Health Statistical Yearbook. 2022: Beijing.

46. Li, M., et al., The effect of pairing assistance under medical alliance policy on healthcare utilization for patients with chronic diseases in rural China. 2023.

47. World Health Organization, Innovative care for chronic conditions : building blocks for actions : global report. 2002, World Health Organization: Geneva.

48. Asch, S.M., et al., Who is at greatest risk for receiving poor-quality health care? N Engl J Med, 2006. 354(11): p. 1147–1156.

49. Jackson, J.L., A. Farkas, and C. Scholcoff, Does Provider Gender Affect the Quality of Primary Care? Journal of General Internal Medicine, 2020. 35(7): p. 2094–2098.

50. Peabody, J.W., et al., Comparison of vignettes, standardized patients, and chart abstraction: a prospective validation study of 3 methods for measuring quality. JAMA, 2000. 283(13): p. 1715–1722.

